# Weight Loss-Independent Changes in Human Growth Hormone During Water-Only Fasting: A Secondary Evaluation of a Randomized Controlled Trial

**DOI:** 10.1101/2024.05.28.24308055

**Authors:** Benjamin D. Horne, Jeffrey L. Anderson, Heidi T. May, Tami L. Bair, Viet T. Le, Leslie Iverson, Kirk U. Knowlton, Joseph B. Muhlestein

## Abstract

**Introduction:** Water-only fasting for one day or more may provide health benefits independent of weight loss. Human growth hormone (HGH) may play a key role in multiple fasting-triggered mechanisms. Whether HGH changes during fasting are independent of weight loss and how basal HGH and HGH change relate to other fasting-induced changes are unknown.

**Methods:** Apparently healthy individuals (N=30) were randomized by Latin square to begin two days with either 24-hour water-only fasting or a control of 24-hour *ad libitum* eating. On day 2, subjects were crossed over to control (if day 1 was fasting) or fasting (if they ate on day 1). HGH, weight, and other parameters were measured at baseline and at the end of the first and second days.

**Results:** Baseline HGH had median 0.50 ng/mL for females (n=20) and 0.04 ng/mL for males (n=10), and correlated inversely with weight, glucose, insulin, and triglycerides and positively with changes in insulin and HOMA-IR. The 24-hour fasting-induced HGH change was uncorrelated with weight loss (r= 0.01, p=0.98), but correlated with changes in glucose, HGB, and IGF-1. The percent increase in HGH was greater (p<0.001) for lower (females ≤0.15 ng/mL, males ≤0.05 ng/mL) vs. higher baseline HGH (median: 1,225% vs. 50.3%, respectively). Subjects with lower baseline HGH had a trend to greater reduction of HOMA-IR (median: −6.15 vs. −1.35 for lower vs. higher HGH, respectively, p=0.08).

**Conclusions:** Fasting increased HGH and the HGH changes were independent of weight loss. Basal HGH and fasting-induced HGH changes correlated inversely with cardiometabolic risk factors.

## Introduction

Intermittent fasting reportedly triggers various health benefits, the most studied of which is weight loss.(1) Fasting may also improve major cardiovascular and metabolic risk factors for chronic disease such as blood pressure, cholesterol levels, and glucose concentrations,(1) and activate other possible benefits through various mechanisms. Fasting triggers health benefits through multiple pathways, including weight loss that leads to consequent health improvements,(2–6) chronobiological effects that may normalize circadian rhythms of glucose metabolism irrespective of weight loss,(7,8) and other mechanisms that are not dependent on weight change.(9–14) Crucially, fasting for durations longer than 20 hours may induce a multitude of weight loss-independent benefits.(9–15)

Some weight loss-independent benefits of fasting involve a metabolic transition in the human body from the use of glucose for energy to the use of fatty acid-derived ketones.(9,10,12,13,15) Use of ketones for energy depends primarily on the duration of complete or near-complete restriction of energy intake.(9,10,12,13,15) The consequences of ketone use may include the reduction of insulin resistance.(14) Other fasting duration-dependent mechanisms may include activating and enhancing autophagy,(16–19) mitophagy,(20–23) and mitochondriogenesis,(23) regulating energy expenditure,(24,25) the immune system (17), and inflammation,(17,26,27) and engineering a healthy microbiome.(12,28) Further duration-driven benefits may ameliorate heart failure (HF) risk by triggering natriuresis,(11,29,30) increasing hemoglobin without hemoconcentration,(11,30) and (based on an animal model) protecting the heart from stress while inducing stronger myocardial contraction.(31)

Finally, the HGH/insulin-like growth factor-1 (IGF-1) axis and especially reduced HGH production is involved in HF development and progression.(32–34) Prolonged water-only fasting substantially increases endogenous HGH during the fasting period.(11,35–37) HGH regulates a broad array of metabolic actions in adults. It exerts direct impacts on protein conservation during episodes of fasting, including the stimulation of protein synthesis and the sparing of lean muscle mass.(35,36) HGH also directly regulates IGF-1 and insulin, and can activate other anabolic agents.(36) In adipose tissue, HGH has a catabolic effect where it metabolizes triglycerides into free fatty acids.(38) As noted above, this triggering of increased concentrations of circulating ketones is one of the central pathways of weight loss-independent benefits from fasting. Further, HGH may reduce the risk of developing HF and, in people with diagnosed HF, HGH may prevent further degradation in myocardial function.(32–34) This study evaluated whether changes in HGH due to 24-hour water-only fasting correlate with change in weight to determine whether these changes are independent and secondarily evaluated whether basal HGH or fasting-induced change in HGH concentrations are associated with baseline cardiovascular and metabolic risk factors or their changes during water-only fasting.

## Methods

### Study Design, Population, and Prior Findings

Previously, a randomized controlled cross-over trial evaluated 30 individuals over two 24-hour periods in which one 24-hour period was a water-only fasting intervention and the other was an *ad libitum* control in which the subjects were allowed to eat any food and consume any beverage but were encouraged to follow their usual diet.(11) The order of the intervention and control periods was randomized 1:1 by Latin square design in which n=16 fasted the first day and ate the second day, while the other n=14 ate the first day and fasted the second day. The cross-over design matched intervention and control periods within each subject for all individual-specific factors. The Intermountain Health Institutional Review Board approved the trial and it was registered at ClinicalTrials.gov (NCT010159760) before any subjects were enrolled (January 28, 2010). Participants were consented at enrollment to participate in the processes and parameters of the original trial as well as long-term blood sample storage and testing for evaluation of subsequently derived novel hypotheses using available study samples and data. This new evaluation of HGH and other parameters was a *post hoc* secondary evaluation of existing data originally collected during the conduct of the trial.

Subject dietary consumption prior to the trial and during the two-day trial participation were not recorded under the instruction for the trial that adherence was intended to reflect real-world practices. In the 30 subjects, as previously reported,(11) ages ranged from 18-70 years, 66.7% were female, and the primary previous findings included marked increases over 24 hours of fasting in HGH and hemoglobin (HGB), significant declines in weight, triglycerides, sodium (i.e., natriuresis) and other circulating parameters included in the basic metabolic profile, and homeostatic model assessment of insulin resistance [HOMA-IR, calculated as: (insulin in mIU/L × glucose in mg/dL)/405] and its components, but no substantial changes in blood pressures, high-sensitivity C-reactive protein (hsCRP), or waist circumference.(11) A subsequent evaluation of metabolomic factors confirmed that substantial increases in concentrations of circulating fatty acids occurred during a 24-hour water-only fast in this population.(12)

### Trial Inclusions and Exclusions

Inclusion criteria for the trial were that subjects had to be free of any deliberate fasting for more than 12 hours per episode during a 1-year period prior to enrollment, had to not routinely diet by skipping meals, and had to not engage in caloric restriction by limiting caloric intake to below 80% of the US FDA recommended daily intake during the prior 2 years.(11) People were excluded if they had a history of stroke, history of myocardial infarction, past or current smoking history, prior diagnosis of peripheral vascular disease, current use of insulin, active receipt of any therapy for cancer, any disorders of the immune system or immunodeficiency, current use of an immunosuppressive agent, or prior solid organ transplant in the preceding year.

### Data Collection

Peripheral blood samples were drawn at baseline (0 hours) in the post-prandial state within 30 minutes after a standard meal, at the 24 hour end of the first day-long period (either the end of the fasting day or the end of the control day, depending on the randomized sequence of the two day-long periods), and at 48 hours (either the end of the control day or the end of the fasting day, respectively).(11) Testing at the end of the fasting day was conducted prior to breaking of the fast. Samples were either tested in the central clinical laboratory of Intermountain Health (for lipid panels, hsCRP, insulin, the basic metabolic profile, and the complete blood count) or in the cardiovascular research laboratory (i.e., HGH and IGF-1). Of note, HGH levels in adults differ by sex (clinical normal range for adult males is 0.01-1.00 ng/mL and for adult females is 0.03-10.0 ng/mL). Demographics, weight, height, and waist circumference were collected or measured using standard protocols in a research clinic by a trained clinical research coordinator and were monitored and verified by a clinician.

### Statistical Considerations

Means with standard deviations (SD), medians with interquartile ranges (IQR), and numbers of subjects with percentages are presented for baseline characteristics, as appropriate for each variable. Differences in baseline characteristics between groups defined by HGH measured at baseline were compared using the chi-square test or Student’s T-test, with the exception of hsCRP that was non-normally distributed and was examined using the non-parametric Mann-Whitney test.

For HGH, weight, and HOMA-IR outcomes, because the distributions of baseline HGH, the change in HGH during fasting, and HOMA-IR measures were non-normally distributed, comparisons of means for outcome assessments used the Mann-Whitney test and correlation statistics used the non-parametric Spearman’s rho (a type of rank correlation coefficient). Medians with IQR and means with SDs are presented for HGH-related measures, while weight-related measures are presented with just means with SDs and HOMA-IR related data are presented with medians and IQRs. Analyses used SPSS v.29.0 (IBM SPSS, Armonk, NY) and p≤0.05 was designated as nominally significant for the primary comparisons of HGH change and weight loss. Other comparisons were not corrected for multiple comparisons but were considered confirmatory of prior data or hypothesis-generating results in need of further evaluation in future studies.

## Results

Baseline HGH for females (n=20) was a median of 0.50 ng/mL and interquartile range (IQR): 0.09 to 1.19 ng/mL (mean 1.26±1.89 ng/mL, minimum 0.03 ng/mL, maximum 5.83 ng/mL), while baseline HGH for males (n=10) was median 0.04 ng/mL and IQR: 0.04 to 0.29 ng/mL (mean 0.23±0.40 ng/mL, minimum 0.02 ng/mL, maximum 1.26 ng/mL). Baseline weight averaged 72.5±15.3 kg (median: 71.4 kg, IQR: 61.9 to 80.6 kg) for females and 99.3±31.2 kg (median: 86.9 kg, IQR: 74.4 to 136.1 kg) for males. Age averaged 44 years; other baseline characteristics are shown in Table 1.

**Table 1.** Baseline characteristics of the study population.

| Characteristic | Overall | Lower HGH | Higher HGH | p-value |
| --- | --- | --- | --- | --- |
|  |  | at Baseline* | at Baseline* |  |
| Sample size | N=30 | n=15 | n=15 | ----- |
| Age (years) | 43.6±13.5 | 41.2±13.4 | 46.0±13.6 | 0.34 |
| Sex (female) | 20 (66.7%) | 8 (53.3%) | 12 (80.0%) | 0.12 |
| Weight (kg) (n=27) | 81.5±24.9 | 84.3±26.8 | 78.5±23.3 | 0.55 |
| Height (cm) (n=27) | 170±11 | 170±13 | 170±9 | 0.98 |
| BMI (kg/m<sup>2</sup>) (n=27) | 27.8±6.6 | 28.6±5.5 | 27.1±7.7 | 0.55 |
| Waist Circumference (cm) (n=27) | 86.3±16.2 | 89.2±16.9 | 83.2±15.5 | 0.35 |
| Glucose (mg/dL) | 82.4±17.1 | 82.7±21.4 | 82.1±12.1 | 0.93 |
| Insulin (mIU/L) | 35.5±28.2 | 42.0±32.6 | 28.7±21.6 | 0.21 |
| HOMA-IR | 7.49±6.15 | 8.95±7.05 | 5.93±4.79 | 0.19 |
| Total Cholesterol (mg/dL) | 191±30 | 200±34 | 182±23 | 0.09 |
| LDL-C (mg/dL) | 105±29 | 116±26 | 93±27 | 0.020 |
| HDL-C (mg/dL) | 55.3±15.8 | 49.3±10.8 | 61.2±18.1 | 0.038 |
| Triglycerides (mg/dL) | 143±68 | 173±72 | 114±49 | 0.014 |
| hsCRP (mg/L)† | 1.35 (0.50,3.88) | 1.60 (0.80,5.90) | 1.00 (0.40,2.60) | 0.31 |
| Sodium (mmol/L) | 139.3±1.9 | 139.1±1.4 | 139.5±2.0 | 0.58 |
| Hemoglobin (g/dL) | 14.5±1.3 | 14.7±1.4 | 14.3±1.3 | 0.41 |
| RDW-CV (%) | 13.7±0.8 | 13.8±0.9 | 13.5±0.5 | 0.22 |
| IGF-1 (ng/mL) | 200±89 | 208±68 | 192±110 | 0.66 |
| HGH (ng/mL)<sup>†</sup> | 0.18 (0.88,1.27) | 0.05 (0.04,0.09) | 0.63 (0.52,3.56) | -----‡ |

\*Thresholds were ≤0.15 ng/mL for females and ≤0.05 ng/mL for males for lower baseline HGH and, for higher baseline HGH, were >0.15 ng/mL for females and >0.05 ng/mL for males; <sup>†</sup>Data are median (interquartile range); <sup>‡</sup>The concentrations of baseline HGH were statistically significantly different by design.
BMI: body mass index; HDL-C: high-density lipoprotein cholesterol; HGH: human growth hormone; hsCRP: high-sensitivity C-reactive protein; HOMA-IR: homeostatic model assessment of insulin resistance; IGF-1: insulin-like growth factor-1; LDL-C: low-density lipoprotein cholesterol; RDW-CV: red cell distribution width-coefficient of variation

Baseline HGH was weakly inversely correlated (Table 2) with weight (r= −0.25, p=0.023, Figure 1A), waist circumference, glucose, insulin, and HOMA-IR, was moderately inversely correlated with triglyceride levels, and was positively correlated with HDL-C, change in insulin during fasting, and change in HOMA-IR during fasting. The 24-hour HGH change during fasting did not correlate with fasting-induced weight loss (r= 0.01, p=0.98, Figure 1B) and was also not correlated with baseline HGH (r= −0.15, p=0.45) or baseline weight (r= −0.23, p=0.26), but did correlate with fasting-induced changes in glucose, HGB, and IGF-1 (Table 2). Relative changes in HGH during fasting (normalized to baseline HGH) were correlated with its denominator (i.e., baseline HGH) with r= −0.74 (p<0.001) and numerator (i.e., absolute HGH change) with r=0.70 (p<0.001), along with baseline HDL-C (r= −0.37, p=0.042), but not with baseline weight (r= −0.01, p=0.95) or fasting-induced weight loss (r=0.07, p=0.72).

**Figure 1.**
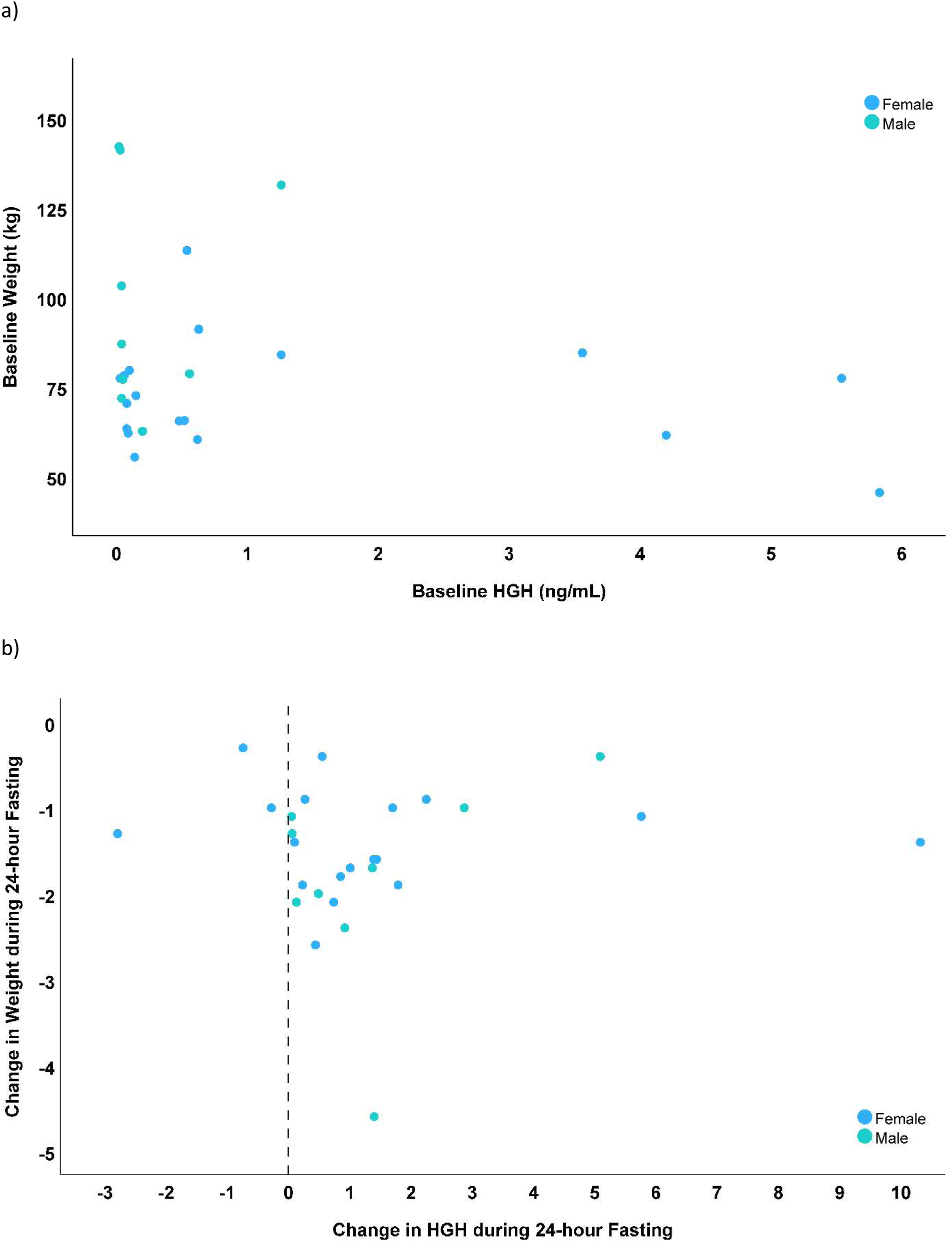
Scatterplots comparing: A) baseline HGH to baseline weight (r= −0.25), and B) fasting-induced changes in HGH to weight loss during fasting (r=0.01).

**Table 2.**
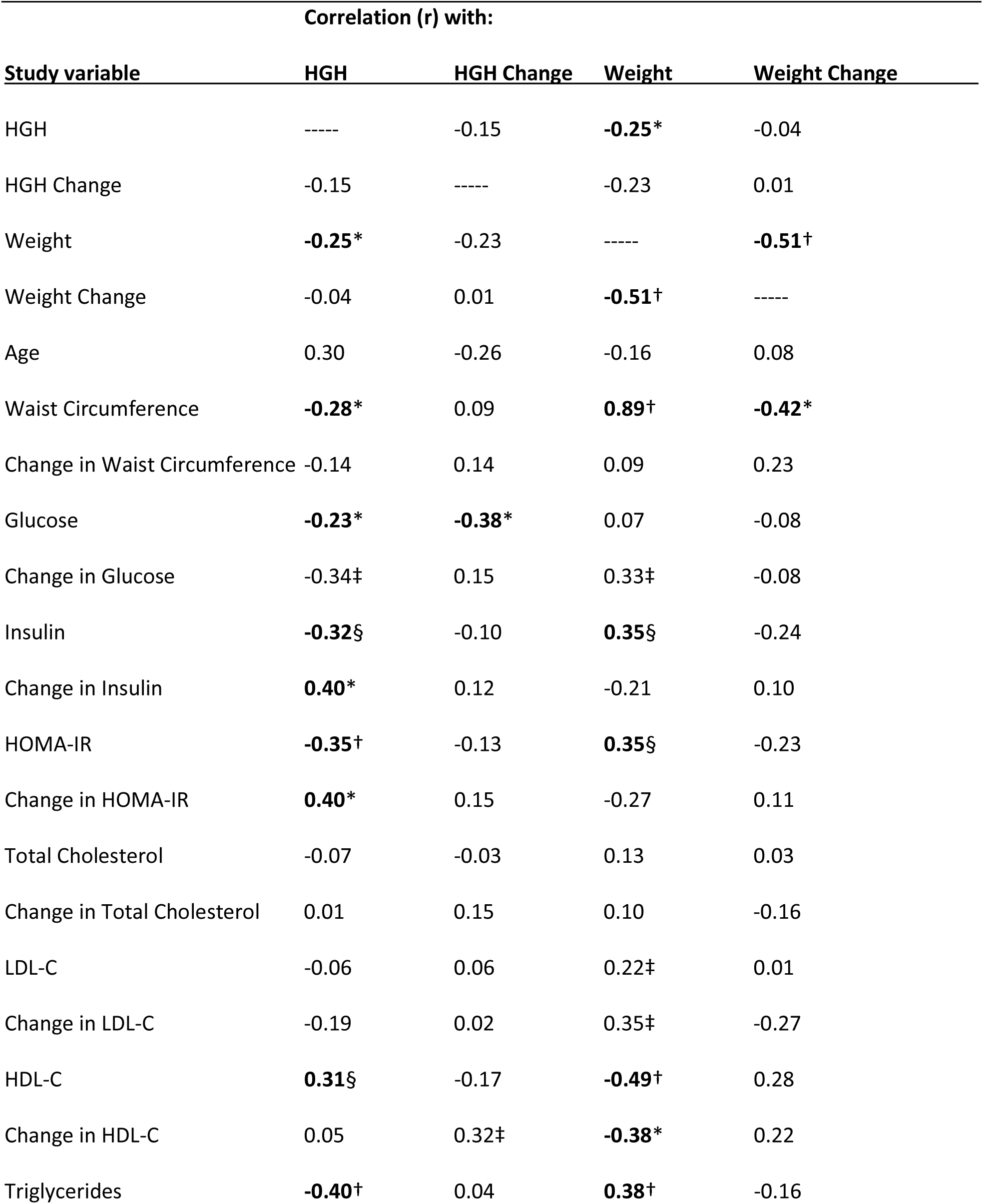
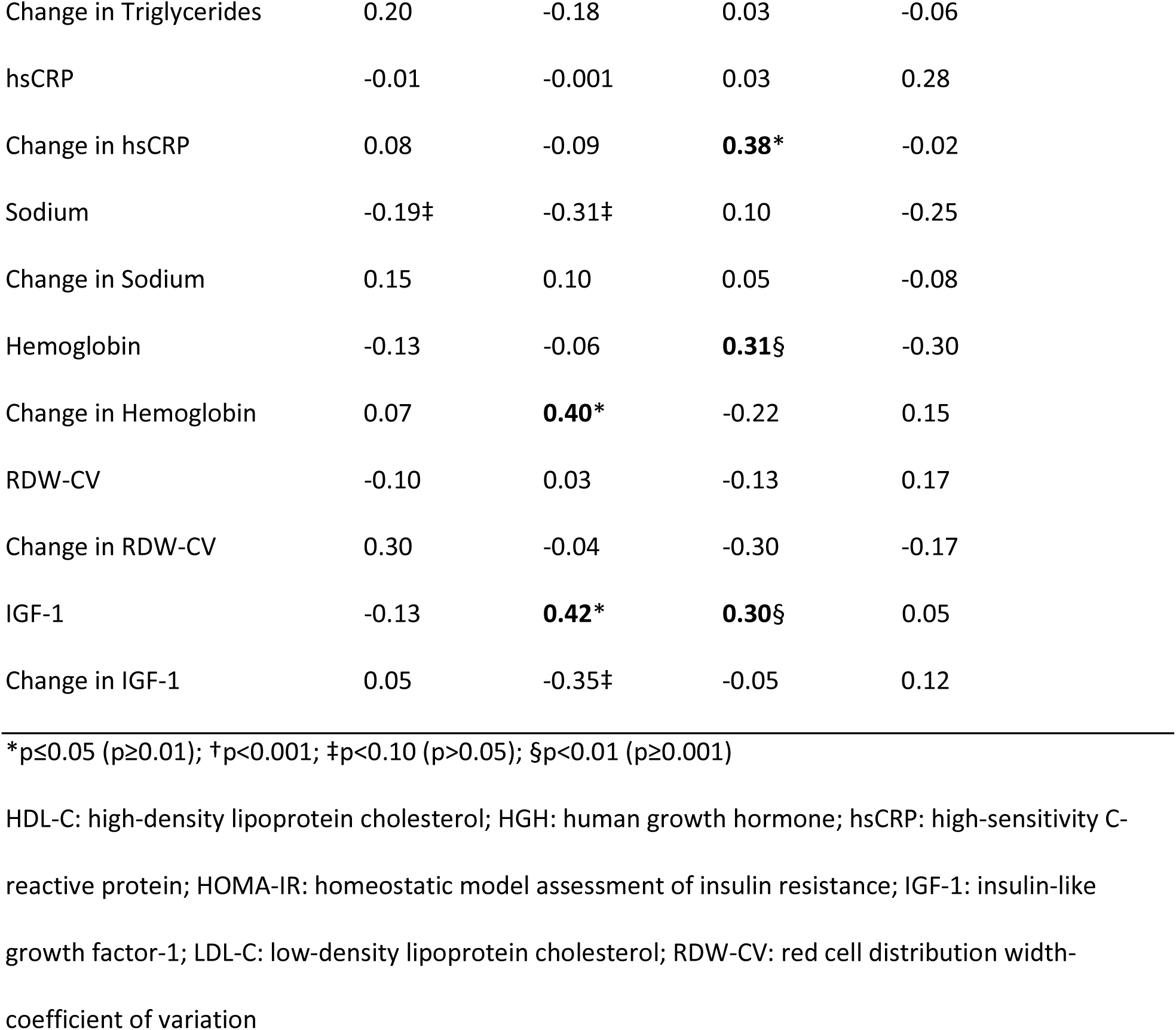
Correlation coefficients of baseline HGH and weight and changes in HGH and weight during 24-hour water-only fasting compared to each other and to other cardiovascular and metabolic risk factors.

| Study variable | Correlation (r) with: |  |  |  |
| --- | --- | --- | --- | --- |
|  | HGH | HGH Change | Weight | Weight Change |
| HGH | ----- | -0.15 | **-0.25\*** | -0.04 |
| HGH Change | -0.15 | ----- | -0.23 | 0.01 |
| Weight | **-0.25\*** | -0.23 | ----- | **-0.51†** |
| Weight Change | -0.04 | 0.01 | **-0.51†** | ----- |
| Age | 0.30 | -0.26 | -0.16 | 0.08 |
| Waist Circumference | **-0.28\*** | 0.09 | **0.89†** | **-0.42\*** |
| Change in Waist Circumference | -0.14 | 0.14 | 0.09 | 0.23 |
| Glucose | **-0.23\*** | **-0.38\*** | 0.07 | -0.08 |
| Change in Glucose | -0.34‡ | 0.15 | 0.33‡ | -0.08 |
| Insulin | **-0.32§** | -0.10 | **0.35§** | -0.24 |
| Change in Insulin | **0.40\*** | 0.12 | -0.21 | 0.10 |
| HOMA-IR | **-0.35†** | -0.13 | **0.35§** | -0.23 |
| Change in HOMA-IR | **0.40\*** | 0.15 | -0.27 | 0.11 |
| Total Cholesterol | -0.07 | -0.03 | 0.13 | 0.03 |
| Change in Total Cholesterol | 0.01 | 0.15 | 0.10 | -0.16 |
| LDL-C | -0.06 | 0.06 | 0.22‡ | 0.01 |
| Change in LDL-C | -0.19 | 0.02 | 0.35‡ | -0.27 |
| HDL-C | **0.31§** | -0.17 | **-0.49†** | 0.28 |
| Change in HDL-C | 0.05 | 0.32‡ | **-0.38\*** | 0.22 |
| Triglycerides | **-0.40†** | 0.04 | **0.38†** | -0.16 |
| Change in Triglycerides | 0.20 | -0.18 | 0.03 | -0.06 |
| hsCRP | -0.01 | -0.001 | 0.03 | 0.28 |
| Change in hsCRP | 0.08 | -0.09 | **0.38\*** | -0.02 |
| Sodium | -0.19‡ | -0.31‡ | 0.10 | -0.25 |
| Change in Sodium | 0.15 | 0.10 | 0.05 | -0.08 |
| Hemoglobin | -0.13 | -0.06 | **0.31§** | -0.30 |
| Change in Hemoglobin | 0.07 | **0.40\*** | -0.22 | 0.15 |
| RDW-CV | -0.10 | 0.03 | -0.13 | 0.17 |
| Change in RDW-CV | 0.30 | -0.04 | -0.30 | -0.17 |
| IGF-1 | -0.13 | **0.42\*** | **0.30§** | 0.05 |
| Change in IGF-1 | 0.05 | -0.35‡ | -0.05 | 0.12 |

\*p≤0.05 (p≥0.01); †p<0.001; ‡p<0.10 (p>0.05); §p<0.01 (p≥0.001)
HDL-C: high-density lipoprotein cholesterol; HGH: human growth hormone; hsCRP: high-sensitivity C-reactive protein; HOMA-IR: homeostatic model assessment of insulin resistance; IGF-1: insulin-like growth factor-1; LDL-C: low-density lipoprotein cholesterol; RDW-CV: red cell distribution width-coefficient of variation

Two categories of baseline HGH were created using sex-specific thresholds for HGH values and ensuring approximately similar sample sizes, with “lower baseline HGH” defined as HGH ≤0.15 ng/mL for females and ≤0.05 ng/mL for males, and “higher baseline HGH” being HGH >0.15 ng/mL for females and >0.05 ng/mL for males. These separated the population into two distinct groups at p<0.001 [mean HGH: 0.07±0.04 ng/mL (median: 0.05 ng/mL) for lower HGH (n=15) vs. 1.77±1.97 ng/mL (median: 0.63 ng/mL) for higher HGH (n=15)], and results were similarly different for females and males (Figure 2A). No difference was found between HGH categories for baseline weight (mean weight: 84.3±26.8 kg for lower HGH group vs. 78.5±23.3 kg for higher HGH group, p=0.65), as was the case for both females and males (Figure 2B). Interestingly, as the correlation results suggested for baseline HGH and HOMA-IR (Table 2), a trend (p=0.08) was found for greater reduction of HOMA-IR (median: −6.15, IQR: −10.01 to −1.03) during fasting in subjects with lower baseline HGH compared to those with higher baseline HGH (median HOMA-IR change: −1.35, IQR: −4.45 to −0.61). This was due to a trend to a potentially greater reduction in insulin during fasting (median: −12.9 mIU/L vs. −4.8 mIU/L, p=0.13) and not glucose change (median: −2.0 mg/dL vs. −9.0 mg/dL, p=0.11) for lower and higher baseline HGH categories, respectively.

**Figure 2.**
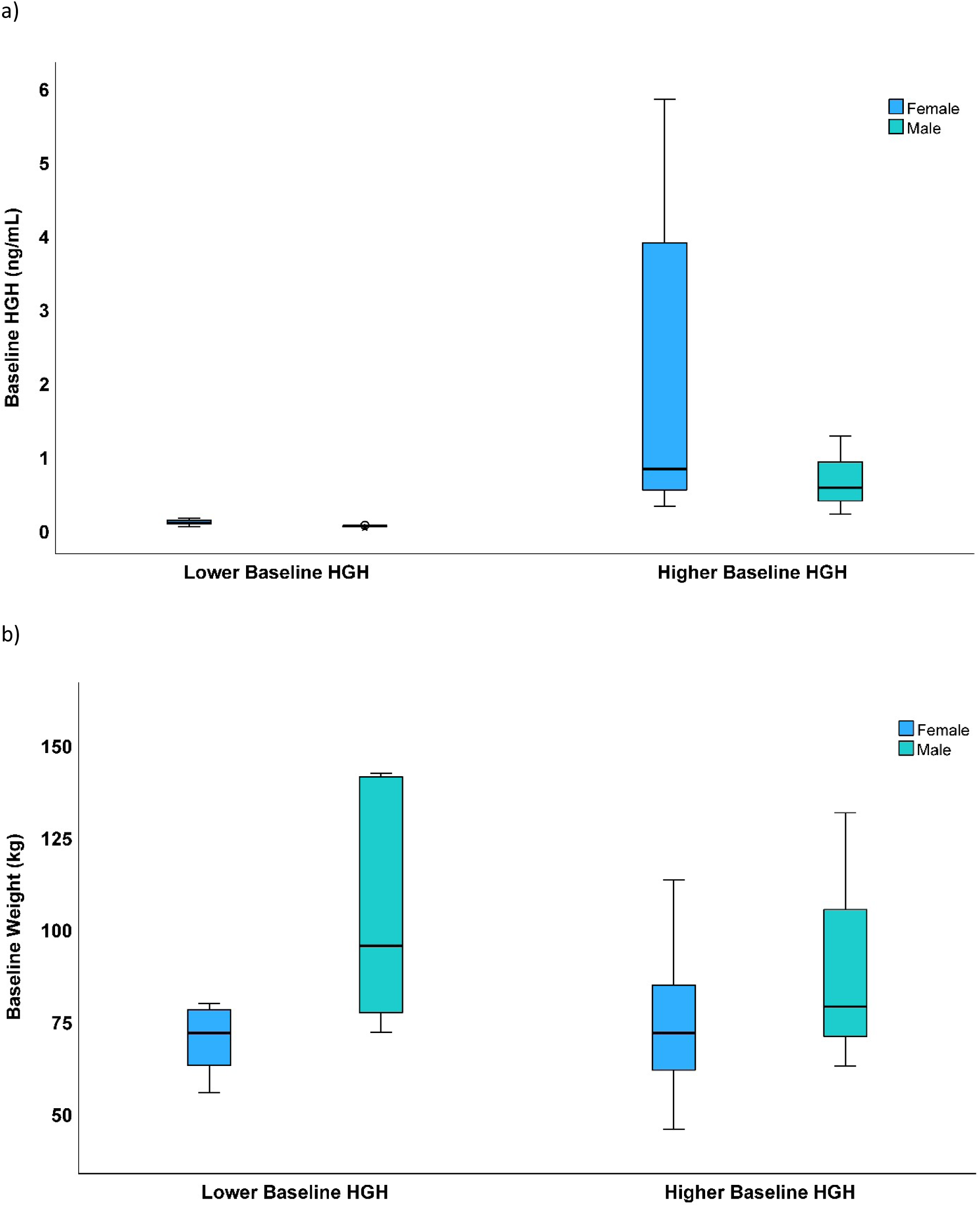
Boxplots displaying sex-specific baseline levels of: A) HGH, and B) weight stratified by baseline HGH categories (Lower HGH: females, ≤0.15 ng/mL; males, ≤0.05 ng/mL. Higher HGH: females, >0.15 ng/mL; males, >0.05 ng/mL). A) For females, the lower HGH category (n=8) had median HGH of 0.09 ng/mL (mean 0.09±0.04 ng/mL, range: 0.03-0.15 ng/mL), and the higher HGH category (n=12) had median HGH 0.81 ng/mL (mean: 2.04±2.12 ng/mL, range: 0.31-5.83 ng/mL). For males, the lower HGH category (n=7) had median HGH 0.04 ng/mL (mean: 0.04±0.01 ng/mL, range: 0.02-0.05 ng/mL) and the higher HGH category (n=3) had median HGH 0.54 ng/mL (mean: 0.67±0.54 ng/mL, range: 0.20-1.26 ng/mL). B) For females, mean baseline weight was 69.8±8.8 kg in the lower HGH category and 74.8±19.2 kg in the higher HGH category (p=0.63), while for males the mean baseline weight was 103.6±31.2 kg for lower HGH and 90.8±35.9 kg for higher HGH (p=0.44). In each boxplot, the bold line inside a box is the median, the upper and lower limits of the box are its 25^th^ and 75^th^ percentiles, and the whiskers above and below are 1.5 times the height of the box.

During the 24-hour fast, the relative increase in HGH (calculated as the percent change during fasting divided by baseline HGH) was greater (p<0.001) in subjects with lower baseline HGH (as noted above, ≤0.15 ng/mL for females and ≤0.05 ng/mL for males), with relative HGH change of median 1,225% (IQR: 450% to 5,740%, mean: 3,930% ±5,656%, minimum 125%, maximum 20,000%). For those with higher baseline HGH, the relative HGH change was median 50.3% (IQR: −47.9% to 274%, mean: 249% ±651%, minimum −58.7%, maximum 2,545%). Sex-specific changes in HGH were similarly larger in the category with lower baseline HGH compared to those with higher baseline HGH (Figure 3A). Relative change in HGH was not different between subjects stratified by baseline weight (median relative HGH change: 274% vs. 375% in subjects with baseline weight below vs. above the sex-specific median, respectively), and results were similarly not different by baseline weight strata when analyzed separately for each sex (Figure 3B).

**Figure 3.**
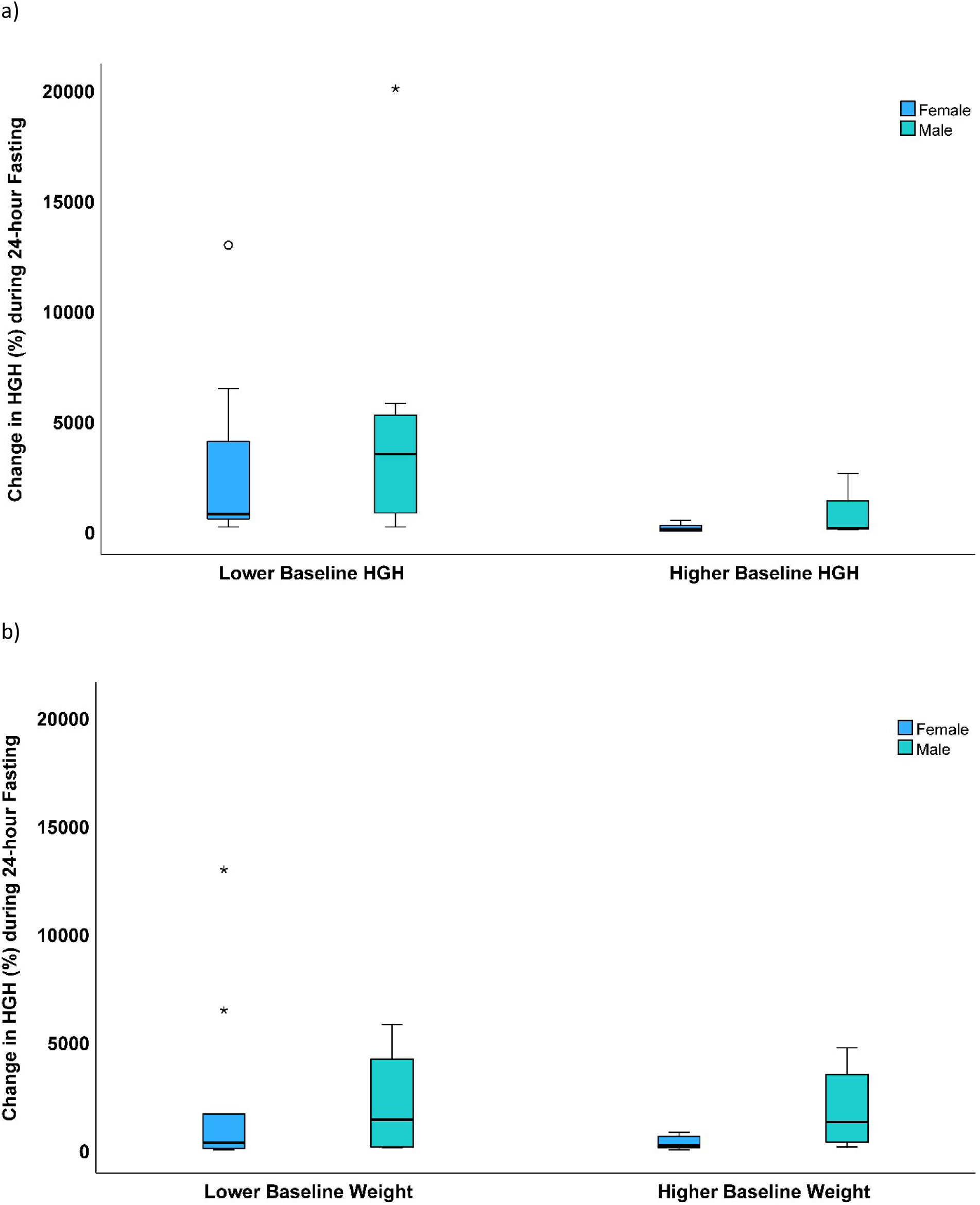
A) Sex-specific boxplots showing the relative increase in HGH during fasting in categories defined by sex-specific thresholds of baseline HGH (p<0.001 for lower vs. higher baseline HGH where lower HGH was ≤0.15 ng/mL for females and ≤0.05 ng/mL for males, and higher HGH was >0.15 ng/mL for females and >0.05 ng/mL for males). Females (n=8) had a median HGH change of 720% (IQR: 475% to 5,202%, mean: 2,934% ±4,514%) in the lower baseline HGH category, while in the higher baseline HGH category females (n=12) had a median HGH change of 38.1% (IQR: −49.8% to 240%, mean: 91% ±162%). For males (p=0.09), the HGH change was a median of 3,425% (IQR: 300% to 5,740%, mean: 5,069% ±6,931%) in the lower baseline HGH category (n=7), while in the higher baseline HGH category males (n=3) had median HGH change of 73.0% (IQR 23.2% to 2,545%, mean: 880% ±1,442%). In each boxplot, the bold line inside a box is the median, the upper and lower limits of the box are its 25^th^ and 75^th^ percentiles, the whiskers above and below are 1.5 times the height of the box, open circles are outliers >1.5 times the height of the box, and asterisks are outliers >3.0 times the height of the box. B) Sex-specific boxplots showing the relative increase in HGH during fasting in categories defined by sex-specific thresholds of baseline weight (Lower baseline weight for females was ≤70 kg and for males was ≤79 kg, while higher baseline weight for females was >70 kg and for males was >79 kg). Median HGH increase in females of 274% (IQR: −13.8% to 4,004%, mean: 2,390% ±4,454%) for those with lower baseline weight (n=9), compared to a median of 137% (IQR: 38.1% to 612%, mean: 296% ±314%) for females (n=9) with higher baseline weight (p=0.76). For males, the relative HGH change was a median of 1,335% (IQR: 48.7% to 4,941%, mean: 2,108% ±2,687%) in the lower baseline weight category (n=4), while in the higher baseline weight category males (n=5) had median HGH change of 1,225% (IQR: 187% to 4,046%, mean: 1,938% ±2,020%) (p=0.81).

## Discussion

### HGH, Fasting, and Cardiometabolic Disease

In addition to stimulating protein synthesis, sparing lean muscle, and regulating IGF-1 and insulin,(35,36) HGH triggers a catabolic effect in adipose tissue that transforms stored triglycerides into fatty acids.(38) This HGH action increases the concentration of circulating fatty acids that are subsequently converted in the liver into beta-hydroxybutyrate and other ketones.(9,10,13) When ketone exposure is of limited duration, their use in myocardial cells of an animal model promoted stronger contraction than when glucose was utilized for energy and they may thus interdict the deterioration of myocardial strength.(31) Ketones produced similar results during *in vitro* examination of human myocardial cells derived from failing hearts, with contractile dysfunction improved when additional substrates were added.(39)

The length of elevated HGH exposure is important. Long-term deficiency of HGH is associated with greater severity of cardiovascular disease and poor prognosis after heart disease diagnosis.(32–34) More generally, decreased expression of HGH/IGF-1 axis is linked to advanced age and greater adiposity.(40) In apparently healthy males aged >60 years who had chronically low plasma IGF-1, administration of HGH for 6 months increased plasma IGF-1 from <350 U/L to 500-1500 U/L and increased lean body mass by 8.8% and skin thickness by 7.1%,(40) while adipose tissue was reduced by 14.4%.(40) Beyond disease, HGH administration in athletes reduced fat mass and increased lean body mass and sprint capacity.(41) In contrast, chronically-elevated HGH is linked to poor HF outcomes, including increased insulin resistance.(33,38)

Achieving optimal HGH levels for an optimal duration may, therefore, be optimal for metabolic and cardiovascular health. This intermittent HGH elevation may be realized through HGH replacement therapy or potentially via intermittent or periodic water-only extended fasting.(42) A 40-hour fast increased HGH, reduced free IGF-1, and increased IGFBP-1 in one study where HGH hypersecretion was not simply the result of reduced free IGF-1.(37) In another study, HGH increased substantially by 5-fold for males and 14-fold for females during a 24-hour water-only fast.(11) Additional studies focusing on the effects of starvation in humans also showed that calorie-free fasting increases HGH.(35,36) Although such fasting periods of 24-40 hours of calorie-free time are longer than advocated by the most popular intermittent fasting regimens (e.g., time-restricted eating and twice-per-week modified fasting), they are relatively short periods compared to some fasting regimens that have been studied.(15) Their results suggest that extended fasting need not involve fasting for more than 2 days to activate key weight loss-independent effects of fasting.

While diabetes and coronary heart disease may be prevented or treated by fasting,(1–8) the development and progression of HF also involves various metabolic and cardiovascular pathways,(43) and these may be ameliorated by water-only fasting through its effects on HGH and other pathways.(32–34,39,42) Fasting induces natriuresis, causing selective excretion of sodium while preserving blood volume.(11,29,30) Natriuresis reduces blood pressure, cardiac load, and risk of poor HF outcomes, and periodic fasting is associated with lower B-type natriuretic peptide.(44) A 24-hour fast also increases hemoglobin without hemoconcentration, increasing oxygen carrying capacity and reducing anemia.(11,30) In animal models, fasting activated autophagy and reversed cardiomyopathy,(45) and BHB protected the heart against fasting-induced stress.(31) In addition to neurohormonal influences, myocardial contractility is heavily influenced by dysregulation of metabolic pathways involving diminished fatty acid oxidation, reduced ketone use, and dysfunctional mitochondria.(46) Given the HGH effects of water-only fasting noted above and described in the current study, these various mechanisms that are triggered by fasting may reduce HF risk independently of weight loss.(44)

### Fasting-Induced HGH Elevation and Weight Loss

A recent study of 7-day water-only fasting reported substantial changes in more than 1,000 plasma proteins where the changes began only after more than 3 days of fasting.(15) Crucially, that week-long fast caused approximately 10-fold more changes in plasma proteins that correlated with changes in beta-hydroxybutyrate but not with weight loss, compared to the protein changes that did correlate with weight changes and not with changes in beta-hydroxybutyrate.(15) This study expands on those findings by showing specifically that elevations of HGH during fasting are independent of weight loss and by demonstrating in a larger study population that such changes occurred after just 24 hours of water-only fasting.

In the present study, the short-term (<3 days) elevation of HGH levels was shown through a water-only fasting regimen that was of a sufficiently extended duration for meaningful changes in HGH and other weight loss-independent proteins to occur but that was not so long that most people would be unwilling to attempt it or that HGH would be elevated continuously for a substantial length of time. Herein, thresholds of lower and higher HGH measurements were based on HGH levels achieved during fasting for people who were apparently healthy and had no recent history of routine fasting for extended periods of time. Subjects with lower HGH appeared to experience superior benefit during the fasting period, and one question that arises from these findings is whether the metabolic benefit from repeated episodes of low-dose intermittent fasting over a period of months or years would yield a greater health benefit based on long-term improvements in insulin sensitivity.

This differential effect dependent on basal HGH may in part be based on cardiometabolic risk levels and, therefore, the health needs of the individual. Baseline HGH concentrations herein correlated inversely with multiple cardiac risk factors including glucose, insulin, HOMA-IR, weight, waist circumference, and triglycerides, and it correlated positively with HDL-C. Pre-fasting HGH also positively correlated with changes in insulin and HOMA-IR, suggesting a possible role for HGH as a marker of the personalized level of benefit that will be received from engaging in fasting. Basal HGH could potentially be used to guide the frequency or intensity of fasting that is needed to improve health for certain people with initially low HGH, although this requires further prospective testing. HGH is established to directly regulate insulin levels, and thus should impact HOMA-IR as these data suggest. Furthermore, change in HGH correlated inversely with glucose and positively with change in HGB and baseline IGF-1, showing no connection of HGH change to change in weight but indicating that additional research is necessary. In contrast, weight loss during fasting only correlated with baseline weight and waist circumference, and not with other clinical cardiovascular or metabolic factors.

### Limitations

While this study evaluated data that were prospectively collected by a randomized controlled trial, the evaluation may be limited by the *post hoc* nature of the hypotheses chosen for examination here since they were not part of the original design or purpose of the trial. The trial randomization may not have controlled all factors related to change in HGH, although analyses herein (see Table 1) suggested that minimal differences existed between those with lower and higher baseline HGH. Further exploration of the hypotheses is needed. The pilot nature of the study, given the sample size of 30 subjects, also suggests a need for further evaluation to validate these findings, including in populations with different ethnic and racial characteristics. The short-term single fasting period of one 24-hour period also limits the interpretation of the findings for long-term health outcomes, but it does provide evidence that substantial health changes occur during fasting and may have meaningful mechanistic importance from a human biological perspective.

### Conclusions

Changes in HGH during 24-hour water-only fasting were not correlated with changes in weight, indicating that health benefits realized through HGH elevation during fasting may be achieved even if no weight is lost. Change in HGH during fasting may reflect present cardiometabolic health, with those having greater chronic disease risks experiencing larger increases in HGH during fasting. Long-term studies of repeated episodes of fasting are required to validate these findings. The study also established sex-specific thresholds of baseline HGH in the clinical normal range for use in future studies to examine whether some individuals receive greater benefit from fasting based on their health status or physiological needs. Whether repeated fasting increases basal HGH is unknown. Future research should examine whether people with lower basal HGH receive greater benefits such as reductions in insulin and HOMA-IR compared to people with higher basal HGH and if long-term use of intermittent fasting can increase basal HGH to a level more conducive to long-term homeostasis as reflected by greater healthspan and lifespan.

## Data Availability

The data underlying this article cannot be shared publicly due to United States privacy laws. The data will be shared on reasonable request to the corresponding author.

## References

1. Patikorn C, Roubal K, Veettil SK, Chandran V, Pham T, Lee YY, Giovannucci EL, Varady KA, Chaiyakunapruk N. Intermittent fasting and obesity-related health outcomes: an umbrella review of meta-analyses of randomized controlled trials. JAMA Netw Open 2021;4(12):e2139558.

2. Trepanowski JF, Kroeger CM, Barnosky A, Klempel MC, Bhutani S, Hoddy KK, Gabel K, Freels S, Rigdon J, Rood J, Ravussin E, Varady KA. Effect of alternate-day fasting on weight loss, weight maintenance, and cardioprotection among metabolically healthy obese adults: a randomized clinical trial. JAMA Intern Med 2017;177(7):930–938.

3. Carter S, Clifton PM, Keogh JB. Effect of intermittent compared with continuous energy restricted diet on glycemic control in patients with type 2 diabetes: a randomized noninferiority trial. JAMA Netw Open 2018;1(3):e180756–e180756.

4. Jamshed H, Steger FL, Bryan DR, Richman JS, Warriner AH, Hanick CJ, Martin CK, Salvy SJ, Peterson CM. Effectiveness of early time-restricted eating for weight loss, fat loss, and cardiometabolic health in adults with obesity. A randomized clinical trial. JAMA Intern Med 2022;182(9):953–962.

5. Lin S, Cienfuegos S, Ezpeleta M, Gabel K, Pavlou V, Mulas A, Chakos K, McStay M, Wu J, Tussing-Humphreys L, Alexandria SJ, Sanchez J, Unterman T, Varady KA. Time-restricted eating without calorie counting for weight loss in a racially diverse population. A randomized controlled trial. Ann Intern Med 2023;176(7):885–895.

6. Pavlou V, Cienfuegos S, Lin S, Ezpeleta M, Ready K, Corapi S, Wu J, Lopez J, Gabel K, Tussing-Humphreys L, Oddo VM, Alexandria SJ, Sanchez J, Unterman T, Chow LS, Vidmar AP, Varady KA. Effect of time-restricted eating on weight loss in adults with type 2 diabetes. A randomized controlled trial. JAMA Netw Open 2023;6(10):e2339337.

7. Sutton EF, Beyl R, Early KS, Cefalu WT, Ravussin E, Peterson CM. Early time-restricted feeding improves insulin sensitivity, blood pressure, and oxidative stress even without weight loss in men with prediabetes. Cell Metab 2018;27:1212–1221.e3.

8. Jamshed H, Beyl RA, Della Manna DL, Yang ES, Ravussin E, Peterson CM. Early time-restricted feeding improves 24-hour glucose levels and affects markers of the circadian clock, aging, and autophagy in humans. Nutrients 2019;11(6):1234.

9. Ruderman NB, Aoki TT, Cahill GF Jr. 1976. Gluconeogenesis and its disorders in man. In Gluconeogenesis: Its Regulation in Mammalian Species, ed. Hanson RW, Mehlman MA, pp 515–530. New York: Wiley.

10. Laffel L. Ketone bodies: a review of physiology, pathophysiology and application of monitoring to diabetes. Diabetes Metab Res Rev 1999;15:412–426.

11. Horne BD, Muhlestein JB, Lappé DL, May HT, Carlquist JF, Galenko O, Brunisholz KD, Anderson JL. Randomized cross-over trial of short-term water-only fasting: metabolic and cardiovascular consequences. Nutr Metab Cardiovasc Dis 2013;23:1050–1057.

12. Washburn RL, Cox JE, Muhlestein JB, May HT, Carlquist JF, Le VT, Anderson JL, Horne BD. Pilot study of novel intermittent fasting effects on metabolomics and trimethylamine N-oxide changes during 24-hour water-only fasting in the FEELGOOD Trial. Nutrients 2019;11(2):246.

13. Deru LS, Bikman BT, Davidson LE, Tucker LA, Fellingham G, Bartholomew CL, Yuan HL, Bailey BW. The effects of exercise on β-hydroxybutyrate concentrations over a 36-h fast: A randomized crossover study. Med Sci Sports Exer 2021;53(9):1987–1998.

14. Bartholomew CL, Muhlestein JB, May HT, Le VT, Galenko O, Garrett KD, Brunker C, Hopkins RO, Carlquist JF, Knowlton KU, Anderson JL, Bailey BW, Horne BD. Randomized controlled trial of once-per-week intermittent fasting for health improvement: the WONDERFUL trial. Eur Heart J Open 2021;1(2):oeab026.

15. Pietzner M, Uluvar B, Kolnes KJ, Jeppesen PB, Frivold SV, Skattebo Ø, Johansen EI, Skålhegg BS, Wojtaszewski JFP, Kolnes AJ, Yeo GSH, O’Rahilly S, Jensen J, Langenberg C. Systemic adaptions to extreme caloric restrictions of different durations in humans. Nat Metab 2024;10.1038/s42255-024-01008-9.

16. Alirezaei M, Kemball CC, Flynn CT, Wood MR, Whitton JL, Kiosses WB. Short-term fasting induces profound neuronal autophagy. Autophagy 2010;6(6):702–710.

17. Hannan MA, Rahman MA, Rahman MS, Sohag AAM, Dash R, Hossain KS, Farjana M, Uddin MJ. Intermittent fasting, a possible priming tool for host defense against SARS-CoV-2 infection: Crosstalk among calorie restriction, autophagy and immune response. Immunol Lett 2020;226:38–45.

18. Gnoni M, Beas R, Vásquez-Garagatti R. Is there any role of intermittent fasting in the prevention and improving clinical outcomes of COVID-19? Intersection between inflammation, mTOR pathway, autophagy and calorie restriction. Virus Dis 2021;32:625–634.

19. DiNicolantonio JJ, McCarty M. Autophagy-induced degradation of Notch1, achieved through intermittent fasting, may promote beta cell neogenesis: Implications for reversal of type 2 diabetes. Open Heart 2019;6:e001028.

20. Lettieri-Barbato D, Cannata SM, Casagrande V, Ciriolo MR, Aquilano K. Time-controlled fasting prevents aging-like mitochondrial changes induced by persistent dietary fat overload in skeletal muscle. PLoS One 2018;13(5):e0195912.

21. Madkour MI, El-Serafi AT, Jahrami HA, Sherif NM, Hassan RE, Awadallah S, Faris MAE. Ramadan diurnal intermittent fasting modulates SOD2, TFAM, Nrf2, and sirtuins (SIRT 1, SIRT3) gene expressions in subjects with overweight and obesity. Diabetes Res Clin Pract 2019;155:107801.

22. Zhao Y, Jia M, Chen W, Liu Z. The neuroprotective effects of intermittent fasting on brain aging and neurodegenerative diseases via regulating mitochondrial function. Free Radic Biol Med 2022;182:206–218.

23. Mehrabani S, Bagherniya M, Askari G, Read MI, Sahebkar A. The effect of fasting or calorie restriction on mitophagy induction: a literature review. J Cachexia Sarcopenia Muscle 2020;11:1447–1458.

24. Walton CM, Jacobsen SM, Dallon BW, Saito ER, Bennett SLH, Davidson LE, Thomson DM, Hyldahl RD, Bikman BT. Ketones elicit distinct alterations in adipose mitochondrial bioenergetics. Int J Molec Sci 2020;21:6255.

25. Demine S, Renard P, Arnould T. Mitochondrial uncoupling: a key controller of biological processes in physiology and diseases. Cells 2019;8:795.

26. Han K, Singh K, Rodman MJ, Hassanzadeh S, Wu K, Nguyen A, Huffstutler RD, Seifuddin F, Dagur PK, Saxena A, McCoy JP, Chen J, Biancotto A, Stagliano KER, Teague HL, Mehta NN, Pirooznia M, Sack MN. Fasting-induced FOXO4 blunts human CD4^+^ T helper cell responsiveness. Nat Metab 2021;3:318–326.

27. Han K, Singh K, Rodman MJ, Hassanzadeh S, Baumer Y, Huffstutler RD, Chen J, Candia J, Cheung F, Stagliano KER, Pirooznia M, Powell-Wiley TM, Sack MN. Identification and validation of nutrient state-dependent serum protein mediators of human CD4^+^ T cell responsiveness. Nutrients 2021;13:1492.

28. Maifeld A, Bartolomaeus H, Löber U, Avery EG, Steckhan N, Markó L, Wilck N, Hamad I, Šušnjar U, Mähler A, Hohmann C, Chen CY, Cramer H, Dobos G, Lesker TR, Strowig T, Dechend R, Bzdok D, Kleinewietfeld M, Michalsen A, Müller DN, Forslund SK. Fasting alters the gut microbiome reducing blood pressure and body weight in metabolic syndrome patients. Nat Commun 2021;12(1):1970.

29. Spark RF, Arky RA, Boulter PR, Saudek CD, O’Brian JT. Renin, aldosterone and glucagon in the natriuresis of fasting. N Engl J Med 1975;292:1335e40.

30. Kamel KS, Lin SH, Cheema-Dhadli S, Marliss EB, Haperin ML. Prolonged total fasting: a feast for the integrative physiologist. Kidney Int 1998;53:531–539.

31. Horton JL, Davidson MT, Kurishima C, Vega RB, Powers JC, Matsuura TR, Petuci C, Lewandowski ED, Crawford PA, Muoio DM, Recchia FA, Kelly DP. The failing heart utilizes 3-hydroxybutyrate as a metabolic stress defense. JCI Insight 2019;4(4):e124079.

32. Arcopinto M, Bobbio E, Bossone E, Perrone-Filardi P, Napoli R, Sacca L, Cittadini A. The GH/IGF-1 axis in chronic heart failure. Endocr Metab Immune Disord Drug Targets 2013;13:76–91.

33. Marra AM, Bobbio E, D’Assante R, Salzano A, Arcopinto M, Bossone E, Cittadini A. Growth hormone as biomarker in heart failure. Heart Fail Clinics 2018;14(1):65–74.

34. Cittadini A, Saldamarco L, Marra AM, Arcopinto M, Carlomagno G, Imbriaco M, Del Forno D, Vigorito C, Merola B, Oliviero U, Fazio S, Saccà L. Growth hormone deficiency in patients with chronic heart failure and beneficial effects of its correction. J Clin Endocrinol Metab 2009;94:3329–3336.

35. Nørrelund H, Nair KS, Jørgensen JOL, Christiansen JS, Møller N. The protein-retaining effects of growth hormone during fasting involve inhibition of muscle-protein breakdown. Diabetes 2001;50:96–104.

36. Møller N, Nørrelund H. The role of growth hormone in the regulation of protein metabolism with particular reference to conditions of fasting. Horm Res 2003;59(suppl 1):62–68.

37. Nørrelund H, Frystyk J, Jørgensen JOL, Møller N, Christiansen JS, Ørskov H, Flyvbjerg A. The effect of growth hormone on the insulin-like growth factor system during fasting. J Clin Endocrinol Metab 2003;88(7):3292–3298.

38. Sharma R, Kopchick JJ, Puri V, Sharma VM. Effect of growth hormone on insulin signaling. Molec Cell Endocrinol 2020;518:111038.

39. Vite A, Matsuura TR, Bedi KC, Flam EL, Arany Z, Kelly DP, Margulies KB. Functional impact of alternative metabolic substrates in failing human cardiomyocytes. JACC Basic Transl Sci 2024;9(1):1–15.

40. Rudman D, Feller AG, Nagraj HS, Gergans GA, Lalitha PY, Goldberg AF, Schlenker RA, Cohn L, Rudman IW, Mattson DE. Effects of human growth hormone in men over 60 years old. N Engl J Med 1990;323:1–6.

41. Meinhardt U, Nelson AE, Hansen JL, Birzniece V, Clifford D, Leung KC, Graham K, Ho KKY. The effects of growth hormone on body composition and physical performance in recreational athletes. A randomized trial. Ann Intern Med 2010;152:568–577.

42. Johnson SC. Nutrient sensing, signaling and ageing: The role of IGF-1 and mTOR in ageing and age-related disease. Subcellular Biochemistry (pp. 49–97). 2018, Springer, Singapore.

43. Roger VL. Epidemiology of heart failure. Circ Res 2013;113(6):646–659.

44. Bartholomew CL, Muhlestein JB, Anderson JL, May HT, Knowlton KU, Bair TL, Le VT, Bailey BW, Horne BD. Association of periodic fasting lifestyles with survival and incident major adverse cardiovascular events in patients undergoing cardiac catheterization. Eur J Prev Cardiol 2021;28(16):1774–1781.

45. Ma X, Mani K, Liu H, Kovacs A, Murphy JT, Foroughi L, French BA, Weinheimer CJ, Kraja A, Benjamin IJ, Hill JA, Javaheri A, Diwan A. Transcription factor EB activation rescues advanced αB-crystallin mutation-induced cardiomyopathy by normalizing desmin localization. J Am Heart Assoc 2019;8:e010866.

46. Wende AR, Brahma MK, McGinnis GR, Young ME. Metabolic origins of heart failure. JACC Basic Transl Sci 2017;2(3):297–310.

